# Persistent viral shedding of SARS-CoV-2 in faeces - a rapid review

**DOI:** 10.1101/2020.04.17.20069526

**Authors:** S Gupta, J Parker, S Smits, J Underwood, S Dolwani

## Abstract

**Aim:** In addition to respiratory symptoms, COVID-19 can present with gastrointestinal complaints suggesting possible faeco-oral transmission. The primary aim of this review was to establish the incidence and timing of positive faecal samples for SARS-CoV-2 in patients with COVID-19.

**Methods:** A systematic literature review identified studies describing COVID-19 patients tested for faecal virus. Search terms for Medline included ‘clinical’, ‘faeces’, ‘gastrointestinal secretions’, ‘stool’, ‘COVID-19’, ‘SARS-CoV-2’ and ‘2019-nCoV’. Additional searches were done in AJG, Gastroenterology, Gut, Lancet Gastroenterology and Hepatology, The WHO Database, CEBM, NEJM, social media and the NICE, bioRxiv and medRxiv preprints. Data were extracted concerning the type of test, number and timing of positive samples, incidence of positive faecal tests after negative nasopharyngeal swabs and evidence of viable faecal virus or faeco-oral transmission of the virus.

**Results:** There were 26 relevant articles identified. Combining study results demonstrated that 53·9% of those tested for faecal RNA were positive. Duration of faecal viral shedding ranged from 1 to 33 days after a negative nasopharyngeal swab with one result remaining positive 47 days after onset of symptoms. There is insufficient evidence to suggest that COVID-19 is transmitted via faecally shed virus.

**Conclusion:** There is a high rate of positive PCR tests with persistence of SARS-CoV-2 in faecal samples of patients with COVID-19. Further research is needed to confirm if this virus is viable and the degree of transmission through the faeco-oral route. This may have important implications on isolation, recommended precautions and protective equipment for interventional procedures involving the gastrointestinal tract.

## What does this paper add to the existing literature?

We synthesise all available evidence from multiple sources and clarify the uncertainty around faecal shedding of SARS-CoV-2 virus, its persistence and duration from onset of symptoms, and after negative nasopharyngeal swabs. Evidence for faeco-oral transmission is plausible and demonstrated in one study though its relative contribution to transmission remains unclear.

### Persistent viral shedding of SARS-CoV-2 in faeces – a rapid review

## Introduction

The rapid progression of the COVID-19 pandemic has created significant challenges for the public as well as healthcare professionals around the world. Knowledge regarding virus incubation, transmission and shedding is crucial for the reduction of new cases and protection of health care professionals. Guidance regarding isolation and protective equipment has changed as evidence has increased and developed.

The high incidence of cough and fever in COVID-19 are well established [1]. Gastrointestinal symptoms are also well documented suggesting a potential faeco-oral transmission route [2]. Discharge guidelines for hospitals or declaring a COVID-19 patient recovered in the UK are largely based on time from either symptom onset or positive test depending on severity of illness and discharge destination [3].The European Centre for Disease Prevention and Control (ECDC) on the other hand, has advocated the need for continued self-isolation and hand hygiene measures even 14 days post-discharge based on prolonged viral shedding in faeces and respiratory samples [4]. This evidence may influence the recommended duration of self-isolation, home sanitation practices during isolation and after discharge and the use of protective equipment for procedures involving the gastrointestinal tract. Evidence based recommendations for specialities such as gastroenterology, gastrointestinal endoscopy and gastrointestinal surgery are required where there may be an exposure risk to virus shed in faeces. Despite viral RNA being detected in the air or other surface samples like toilets, it is still unclear whether it is viable to transmit infection through this route [5].

The primary aim of this review is to assess the incidence and timing of positive faecal samples for SARS-CoV-2 in relation to the clinical course of patients with COVID-19. Our secondary aims are to establish the incidence of patients with positive faecal samples after negative respiratory swabs and any evidence to suggest faecal virus transmitted infection.

## Methods

Reports of cases or studies of COVID-19 patients with evidence of the virus in faecal samples were systematically identified and full text articles reviewed for data extraction.

### Literature search

A comprehensive search was undertaken as per the search strategy outlined below for literature which included SARS-CoV-2 virus testing of faeces. Medline was searched to find articles published until 3 April 2020. The defined search terms were created after collaboration between the authors experienced in gastroenterology, colorectal surgery and systematic review. Search terms reflected the aim to identify studies with evidence of faecal COVID-19 and included ‘clinical’, ‘faeces’, ‘gastrointestinal secretions’, ‘stool’, ‘COVID-19’, ‘SARS-CoV-2’ and ‘2019-nCoV’. Additional manual searches to identify the most recent evidence were performed in the American Journal of Gastroenterology (AJG), Gastroenterology, GUT, the Lancet Gastroenterology and Hepatology, the WHO database, the Centre for Evidence Based Medicine (CEBM), the New England Journal of Medicine (NEJM), and the National Institute for Health and Care Excellence (NICE). COVID-19 preprints published until 10 April 2020 on medRxiv and bioRxiv and an independent search on social media (Twitter) by the authors (SS, SD) supplemented more articles. The search strategy used for social media and brief description of the WHO and other databases has been provided in the supplementary methods section.

### Inclusion and exclusion criteria

Articles describing COVID-19 patients who had faecal or stool specimens tested for the virus were included. Considering the knowledge gaps existing for COVID-19 all articles were considered regardless of the number, age or gender of patients or the country of publication. Animal based studies or articles without an available full text were excluded. Foreign language articles were considered but excluded unless the necessary language expertise was available within the research group.

### Study identification

Articles were sorted alphabetically by author name and divided between two reviewers (SG and JP). Abstracts were reviewed and classified by the same two authors through the Rayyan Web Application [6] to identify those for full text review. The same process was used for full text articles and this data was managed through EndNote. Articles were then discussed between the same reviewers to identify the final selection of full text articles. Any conflicts were to be solved by the supervising author if necessary. Reference lists and review articles were cross referenced to identify any further original studies. All articles were categorised and described in a PRISMA flow chart.

### Data extraction

The final data extraction was also carried by the two reviewers (JP and SG) and managed through Microsoft Excel files. The data parameters extracted from the studies are shown in Table 1. The final data was verified by the two reviewers (JP and SG) with conflict resolution as described previously if necessary.

**Table 1.** Data parameters for extraction.

|  |
| --- |
| 1. Study reference<br>2. Country of publication<br>3. Number and type of patients in the study<br>4. Type of sample taken (faecal sample, anal swab, RT-PCR, culture)<br>5. Number of patients having faecal samples tested and number of positive samples<br>6. Timing of positive faecal swab after symptom onset<br>7. Duration of positive faecal specimen after negative nasopharyngeal swab<br>8. Any evidence for viable faecal virus or faeco-oral transmission documented in the study |
| **Table 1 – Data parameters for extraction** |

## Results

Medline searches identified 565 articles and 194 were found through other databases. An overview of the selection process is shown in the PRISMA chart in Figure 1. There were 26 articles [7-32] included in the final analysis. An overview of the patient demographics is summarised in Table 2.

**Figure 1.**
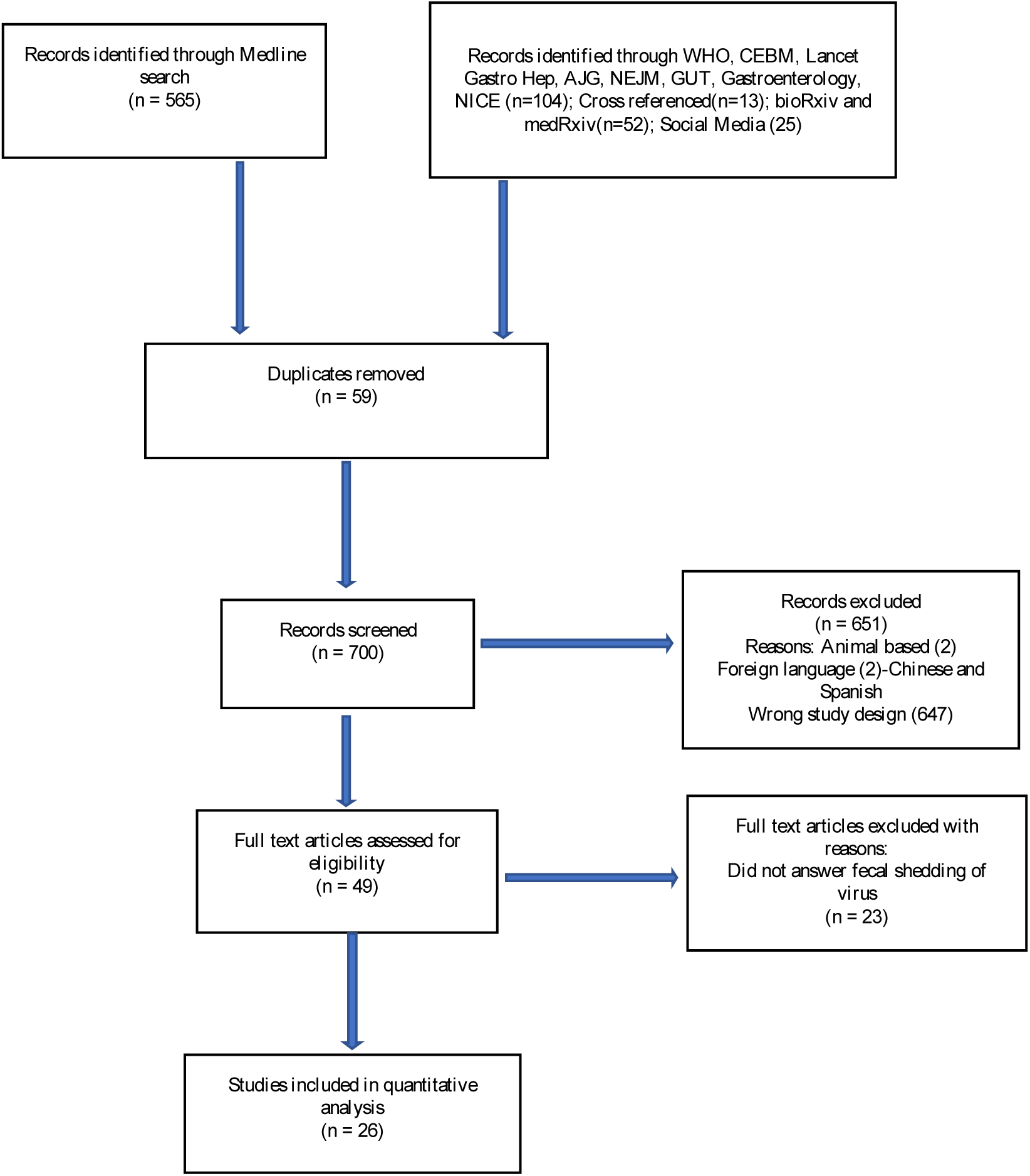
PRISMA flow chart.

**Table 2:** Overview of patient demographics from studies included in the review [7-32].

| Reference | Country | Number of patients in study | Type of patients | Type of sample |
| --- | --- | --- | --- | --- |
| Cai et al. [7] | China | 10 | Children, 3 to 131 months | Faeces |
| Chan et al. [8] | China | 6 | Family cluster (10 to 66 years) | Faeces |
| Chen et al. [9] | China | 1 | Male, 34 years | Faeces |
| Chen et al. [10] | China | 1 | Female, 25 years | Faeces |
| Chen et al. [11] | China | 57 | Unclear | Anal swab |
| Han et al. [12] | China | 206 | Adults | Faeces |
| Holshue et al. [13] | USA | 1 | Male, 35 years | Faeces |
| Kim et al. [14] | Korea | 2 | Adult: male and female | Faeces |
| Kujawski et al. [15] | USA | 12 | Adults | Faeces |
| Lescure et al. [16] | France | 5 | Adults | Faeces |
| Ling et al. [17] | China | 66 | Adults | Faeces |
| Lo et al. [18] | China | 10 | 9 adults, 1 child | Faeces |
| Nicastri et al. [19] | Italy | 1 | Adult, late 20s | Faeces |
| Pan et al. [20] | China | 17 | Laboratory samples | Faeces |
| Peng et al. [21] | China | 9 | Adults | Anal swab |
| Song et al. [22] | China | 1 | Middle aged female | Anal swab |
| Tan et al. [23] | China | 1 | Male, 73 years | Rectal swab |
| Tang et al. [24] | China | 1 | Male, 10 years | Faeces |
| Wang et al. [25] | China | 205 | Adults and children, mean age 44 years | Faeces |
| Wu et al. [26] | China | 74 | Laboratory samples | Faeces |
| Xiao et al. [27] | China | 73 | Children and adults, 10 months to 78 years old | Faeces |
| Xing et al. [28] | China | 3 | Children, 1.5 to 6 years | Faeces |
| Xu et al. [29] | China | 10 | Children, 2 months to 15 years | Rectal swab |
| Zhang et al. [30] | China | 23 | Adults, median age 48 years | Faeces |
| Zhang et al. [31] | China | 14 | Adults, median age 41 years | Faeces |
| Zhang et al. [32] | China | 15 | Laboratory samples | Anal swab |

Most studies were from China (n=20) with two from the USA and one each from Italy, Korea, Vietnam and France. The number of participants recruited in the studies ranged from 1 to 206 with ages ranging from 3 months to 87 years. Sample collection consisted of either faecal samples, anal or rectal swabs. Quantitative Reverse Transcription Polymerase Chain Reaction (RT-PCR) was the test performed on all samples to detect viral RNA. The indication for faecal testing was not specified in most studies. In some the test was done in asymptomatic patients for screening after contact with an infected person or travel history to an infected area. The predominant symptoms of presentation in the studies were persistent cough, fever and breathlessness with fewer patients reporting diarrhoea or vomiting. All studies had information regarding our primary aim of reporting faecal samples for the virus in those with COVID-19. Of these, 16 [7,10,11,14-19,23,24,26–30] provided information on the duration of these tests after symptom onset and evidence of positive faecal samples after symptom recovery, discharge from the hospital or negative nasopharyngeal RT-PCR. The data extraction has been summarised in Table 3 and 4 which are divided based on the number of patients tested for faecal RT-PCR in the study (≤ 10 and >10 respectively) and the detailed combined table is attached as a supplementary results table.

**Table 3:** Overview of data extracted from studies included in the review with ≤ 10 patients tested for faecal virus [7–10,13,14–16,18,19,21–24,28,29].

| Reference | Patients with positive faecal RT-PCR | Timing of positive faecal RT-PCR (from symptom onset unless stated otherwise) | Number of patients with positive faecal RT-PCR and negative NP RT-PCR | Duration of persistent positive faecal RT PCR after negative NP RT-PCR |
| --- | --- | --- | --- | --- |
| Cai et al. [7] | 6 tested, 5 positives (83.3%) | First test at 3-13 days<br>Second test at 18-30 days<br>Positive in all patients on both tests | 5 out of 5 (100%) | Ranged from 11 to 18 days |
| Chan et al. [8] | 4 tested, 0 positive | NA | NA | NA |
| Chen et al. [9] | 1 tested, 0 positive | NA | NA | NA |
| Chen et al. [10] | 1 tested, 1 positive (100%) | Day 11 | 1 out of 1 (100%) | 1 day |
| Holshue et al. [13] | 1 tested, 1 positive (100%) | Day 7 | Not available | Not available |
| Kim et al. [14] | 2 tested, 2 positives (100%) | Ranged from day 8 to 17 | 0 out of 2 | NA |
| Kujawski et al. [15] | 10 tested, 7 positives (70%) | Ranged from day 6 to 18 | 2 out of 7 (28.6%) | Ranged from 4 to 6 days |
| Lescure et al. [16] | 5 tested, 2 positives (40%) | Ranged from day 2 to 13 | 1 out of 2 (50%) | 3 days |
| Lo et al. [18] | 10 tested, 10 positives (100%) | Ranged from day 2 to 19 | 4 out of 10 (40%) | Ranged from 2 to 10 days |
| Nicastri et al. [19] | 1 tested, 1 positive (100%) | Day 3 after admission | 0 out of 1 | NA |
| Peng et al. [21] | 9 tested, 2 positives (22.2%) | Patient 1: day 3<br>Patient 2: unknown | Not available | Not available |
| Song et al. [22] | 1 tested, 0 positive | NA | NA | NA |
| Tan et al. [23] | 1 tested, 1 positive (100%) | Up to day 23 | 1 out of 1 (100%) | 7 days |
| **Tang et al. [24]** | 1 tested, 1 positive (100%) | Ranged from day 17 to 25 after exposure | 1 out of 1 (100%) | 10 days |
| **Xing et al. [28]** | 3 tested, 3 positives (100%) | Patient 1 and 2: day 4<br>Patient 3: day 9 (after discharge) | 3 out of 3 (100%) | 8 and 20 days |
| **Xu et al. [29]** | 10 tested, 8 positives (80%) | Ranged from day 1 to 3 | 8 out of 8 (100%) | Ranged from 3 to 21 days |

NP: Nasopharyngeal; NA: Not applicable; RT-PCR: Reverse Transcriptase Polymerase Chain reaction

**Table 4:** Overview of data extracted from studies included in the review with > 10 patients tested for faecal virus [11,12,17,20,25–27,30–32].

| **Reference** | **Patients with positive faecal RT-PCR** | **Timing of positive faecal RT-PCR (from symptom onset unless stated otherwise)** | **Number of patients with positive faecal RT-PCR and negative NP RT-PCR** | **Duration of persistent positive faecal RT PCR after negative NP RT-PCR** |
| --- | --- | --- | --- | --- |
| **Chen et al. [11]** | 28 tested, 11 positives (39.3%) | Only specify timings in two patients<br>Patient 1: day 13<br>Patient 2: day 10 | 1 out of 2 (50%) | 3 days |
| **Han et al. [12]** | 22 tested, 12 positives (54.5%) | Not available | Not available | Not available |
| **Ling et al. [17]** | 66 tested, 66 positives (100%) | Not available | 43 out of 66 (65%) | Duration to negative NP sample ranged from 6 to 11 days (median 9.5 days)<br>Vs<br>Duration to negative faecal sample ranged from 9 to 16 days (median 11 days)<br>N.B.: 11 patients still had positive faecal RT-PCR at 31 days after admission to convalescence |
| **Pan et al. [20]** | 17 tested, 9 positives (53%) | Ranged from day 0 to 11 | Not available | Not available |
| **Wang et al. [25]** | 153 tested, 44 positives (29%) | Not available | Not available | Not available |
| **Wu et al. [26]** | 74 tested, 41 positives (55%) | Variable | 32 out of 41 (78%) | Faecal sample remained positive for a mean duration of 27.9 days (9.2 days longer than positive respiratory sample)<br>Patient 1: 33 days after -ve nasopharyngeal swab<br>Patient 2: 47 days from symptom onset |
| **Xiao et al. [27]** | 73 tested, 39 positives (53.4%) | Ranged from day 1 to 12 days | 17 out of 39 (23.3%) | Not available |
| **Zhang et al. [30]** | 12 tested, 10 positives (83.3%) | Day 4 | 6 out of 10 (60%) | Median duration of positive NP sample - 10 days<br>Vs<br>Median duration of positive faecal sample - 22 days |
| **Zhang et al. [31]** | 14 tested, 5 positives (35.7%) | Ranged from day 4 to 10 | Not available | Not available |
| **Zhang et al. [32]** | 15 tested, 4 positives (26.7%) | Ranged from day 0 to 5 | Not available | Not available |

NP: Nasopharyngeal; NA: Not applicable; RT-PCR: Reverse Transcriptase Polymerase Chain reaction

A total of 824 patients were included across the studies and 540 were tested for faecal viral RNA [7-32]. Positive faecal RT-PCR tests occurred in 291 (53·9%). The timing of the first positive sample was available in 21 studies and varied from day 0 of symptom onset to day 17. Late positive tests do not necessarily equate to absence of the virus earlier in the illness but may likely reflect the heterogeneity in testing patterns amongst the studies. First stool samples were often reported late after hospital admission [11] or even after discharge [28] while some were analysed from day 1 of hospitalisation or symptom onset [19,20,27,29,32]. There is a similar discrepancy in follow up testing. Some tested until samples were found to be negative [17] while others did not [18,29].

Out of 199 patients who tested positive for faecal viral RNA and were followed up with stool testing, 125 (62·8%) showed persistent shedding of virus in the stool samples after a negative nasopharyngeal swab while in the individual studies it ranged from 23·3% to 100%. The duration for faecal shedding of viral RNA after clearance of respiratory samples ranged from 1 to 33 days and in 1 patient up to 47 days from symptom onset [26].

None of the studies were designed to detect live virus in the faeces except for the study by Wang et al [25]. Out of 153 stool specimens tested in this study, 44 were PCR positive and out of 4 specimens cultured, live virus was detected in 2 [25].

## Discussion

This rapid review demonstrates a high incidence and persistence of positive faecal RT-PCR tests for SARS-CoV-2 after negative nasopharyngeal swabs in patients with COVID-19. This may have important implications regarding measures to prevent the spread of disease, precautions recommended for the public and protective equipment for health professionals performing interventions involving the gastrointestinal tract.

A Chinese review performed by Tian et al. summarised evidence on the importance of identifying gastrointestinal symptoms in addition to the respiratory symptoms of patients with COVID-19 [33]. Despite persistent shedding of SARS-CoV-2 virus in faeces there seems to be no correlation with the presence or severity of gastrointestinal symptoms based on the limited data available. Our review adds to this evidence from China and describes the plausibility of faeco-oral transmission.

Despite this review demonstrating a high incidence of positive tests for virus in the faeces, the absence of evidence to confirm infectivity from this must be emphasised. In order to adequately confirm this, good quality evidence is required to demonstrate infectious virus in faeces and its risk of transmitting disease between individuals. This data may then enable the development of reliable guidelines and recommendations. However, given the rapid development of the pandemic, this will take time and reviews such as this may help guide focussed and valuable research questions for the future. The findings of our review provide a synopsis of the best available evidence regarding SARS-CoV-2 in the faeces at the current time.

Evidence regarding other coronaviruses may be helpful in this context. Similar patterns of virus isolation from stool and faeco-oral transmission were observed for other coronaviruses including SARS-CoV-1 [34]. Bio-aerosol generation of viral particles as a result of toilet flushing, the impact of disinfection on this [35,36] and the persistence of coronaviruses on surfaces has been studied before [37]. Other indirect evidence of microbial exposure and contamination of the operator’s face during endoscopy [38] and laboratory evidence of SARS COV2 infection of the gastrointestinal tract and mechanisms [39,40] add to the evidence for plausibility of transmission.

The risk to health care professionals from patient exposure is well known, specifically in high aerosol generating procedures. Professional societies and investigator groups from countries with experience of managing COVID-19 in the context of gastrointestinal interventions [41,42] highlight the risk to individuals in endoscopy departments and the need for necessary precautions including negative pressure rooms and personal protective equipment for both upper and lower GI procedures. This review supports the importance of these measures given a high prevalence and persistence of SARS-CoV-2 virus in faeces. Isolation of live virus is confirmed only by one study [25] and the proportion of cases that might be transmitted by this route is unclear due to the heterogeneity in case selection and lack of standardisation of study designs and protocols. Environments such as Care Homes may be particularly vulnerable to transmission of infection by this route and recommendations must take into account this evidence to ensure the protection of health and social care providers and the general public in the meantime. Application of this data to the population may be helpful in guiding the recommendations for isolation periods to reduce transmission rates.

### Limitations

Despite finding a high incidence of positive faecal samples for SARS-CoV-2 in the included studies, our review cannot confirm the true population prevalence of positive faecal samples or the rate of false negatives. This is due to the significant variability in study design which is an inherent problem with COVID-19 research at present. This heterogeneity was not formally assessed due to it being a rapid review but can be clearly identified on inspection of the study designs and outcomes. The variability in patient numbers and characteristics, sample timing, sample nature (faecal samples vs. anal or faecal swabs) and follow up testing should be considered when interpreting the reliability of the results. If other studies confirm viable virus in stool, then methods of culture also need to be described and standardised for comparison and replication in other populations. The majority of the included studies are small, heterogenous, retrospective and often did not assess viral shedding in the faeces as their primary aim. At present however this is the only evidence available. There were two foreign language articles excluded due to lack of translation resources. The pre-prints are not peer reviewed and therefore should be trusted with caution.

### Conclusion

The duration of viral shedding in the faeces is mostly reported from 1 to 33 days after a negative nasopharyngeal swab but can continue for up to 47 days after onset of symptoms in patients with COVID-19. These positive samples can occur after negative nasopharyngeal swabs or resolution of patient symptoms. Isolation of live virus in stool specimens of 2 cases in a single study supports the possibility of faeco-oral transmission. Further research is needed to prove if this viral shedding in stool results in a significant proportion of case transmission in the community as well as within care institutions and secondary care. Until further evidence is generated appropriate precautions should be recommended for the protection of healthcare workers and patients.

#### Implications for the Public

1. In addition to strict adherence to hand washing recommendations, home toilet sanitary and disinfection precautions should be taken in the case of isolation or contact with a symptomatic COVID 19 case with or without gastrointestinal symptoms. This statement is based on limited evidence of possible viable faecal virus excretion.
2. These precautions may need to continue for longer than the period of symptoms and the current recommendations for isolation after symptoms cease. This statement is based on limited evidence of the duration after the onset of symptoms that an RT-PCR stool test might still be positive

#### Implications for Healthcare professionals

1. Professional bodies recommendations on protective equipment, endoscopic and surgical procedures for COVID-19 patients should be followed [43–46].
2. The possibility of faeco-oral transmission should be borne in mind with implications for endoscopy and theatre disinfection of surfaces in between procedures.
3. Ward areas for COVID-19 patients and Care homes or similar institutions may need to consider the implications for infection control and disinfection in light of the possibility of faeco-oral transmission
4. Screening processes for patients due to undergo investigational or interventional procedures may need to consider including gastrointestinal symptoms and stool testing in future pre-procedure questionnaires.
5. Healthcare teams managing patients with gastrointestinal symptoms may need to consider the possibility of COVID-19 coexisting with or worsening symptoms of underlying conditions such as Inflammatory Bowel Disease [47].

#### Recommendations for further research

1. Future studies on viral shedding and infectivity of SARS-CoV-2 should consider standardisation of sampling methods in terms of the timing and the type of sample collection, with appropriate precautions for laboratory staff handling these samples until the situation is clearer.
2. Study designs may wish to consider repeat and parallel sampling with nasopharyngeal swabs at defined time points. This may be correlated with symptoms and serology to clarify the effect of neutralising antibodies and viable virus excretion in the stool.
3. Study designs may benefit from testing stool samples from comparable groups. This could include symptomatic, asymptomatic or recovered individuals in and out of family clusters and with or without gastrointestinal symptoms. This may improve our understanding of clinical and public health implications and potential targets for intervention in these settings.

## Data Availability

This is a systematic review and all the data analyzed was from the studies included in the review which have been cited in the manuscript.

## Acknowledgement

We would like to thank Dr SA Roberts for critical review and comments on the manuscript.

## Supplementary methods section

The Mesh terms used for Medline have already been mentioned in the methods section. The search start date was not specified because the terms ‘COVID-19’, ‘SARS-CoV-2’ and ‘2019-nCoV’ used for Medline being novel, themselves limit the start date. The Lancet Gastroenterology and Hepatology was searched for using the terms ‘COVID-19’, ‘SARSCoV-2’ and ‘2019-nCoV’ while in all other databases (WHO, GUT, NICE etc.), and medRxiv and bioRxiv preprints, the COVID-19 articles are mentioned in a separate section. These were manually screened by their titles to find the relevant articles for faecal viral shedding or gastrointestinal symptoms in COVID-19 patients.

Search for Twitter by SS done on 08.04.2020 was as follows:

1. Terms were entered into Twitter search bar

- Terms entered in each separate search were: “shedding faeces cov”, “shedding cov”, “shedding faeces corona” “shedding corona”, “shedding stool cov”, “shedding stool corona”
2. “top” tweets options selected, and all tweets returned in search were read to look for relevant content

- Tweets were read and those that included a link to an article relating to the search term were selected and links to article clicked
- “latest” tweets option selected, and all tweets returned in search were read to look for relevant content
- Tweets were read and those that included a link to an article relating to the search term were selected and links to article clicked

Identified articles were then sent to the core term (SG, JP) and screened for relevance.

Links for accessing the various databases are provided in the following table:

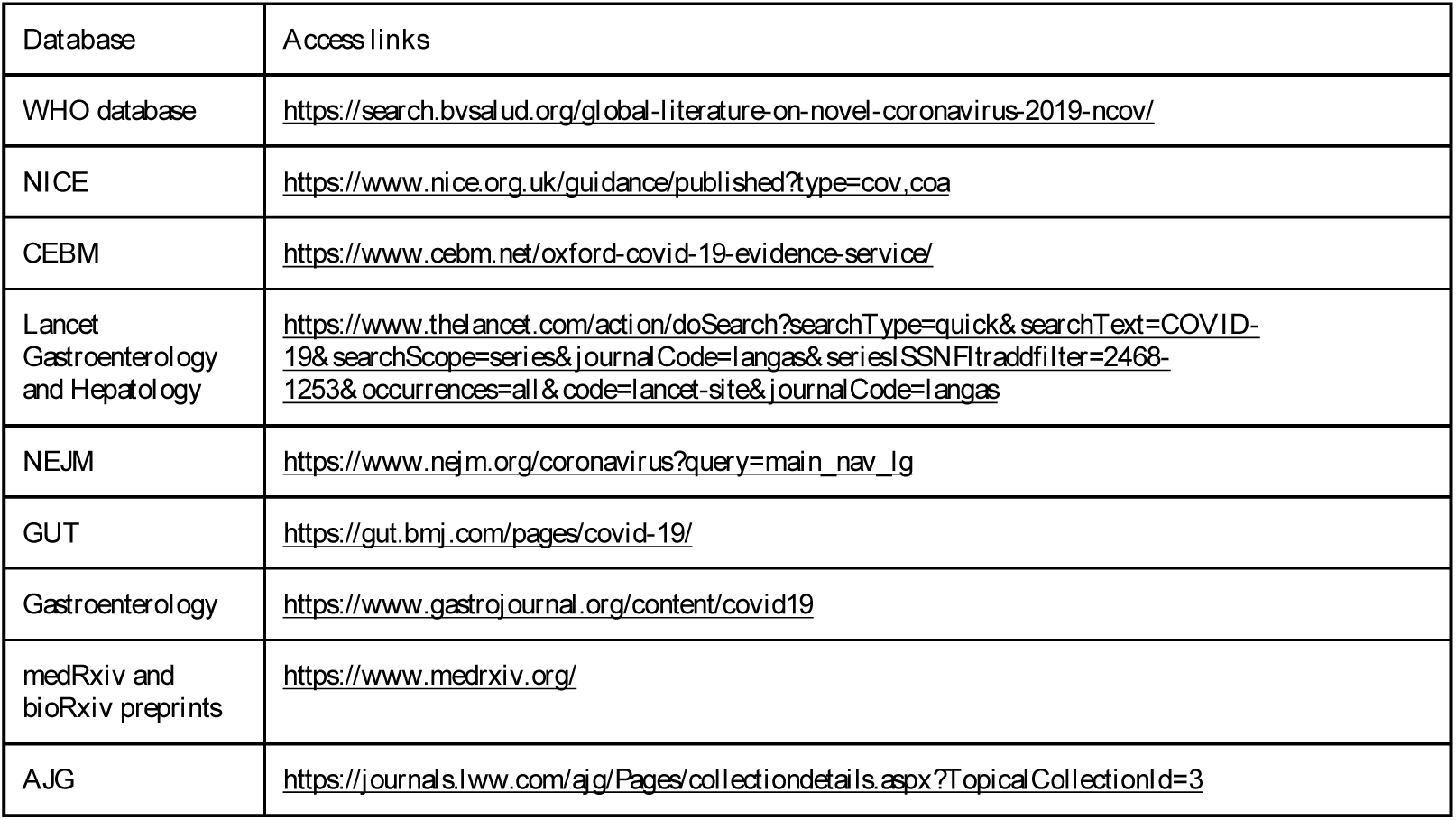

## Supplementary results table

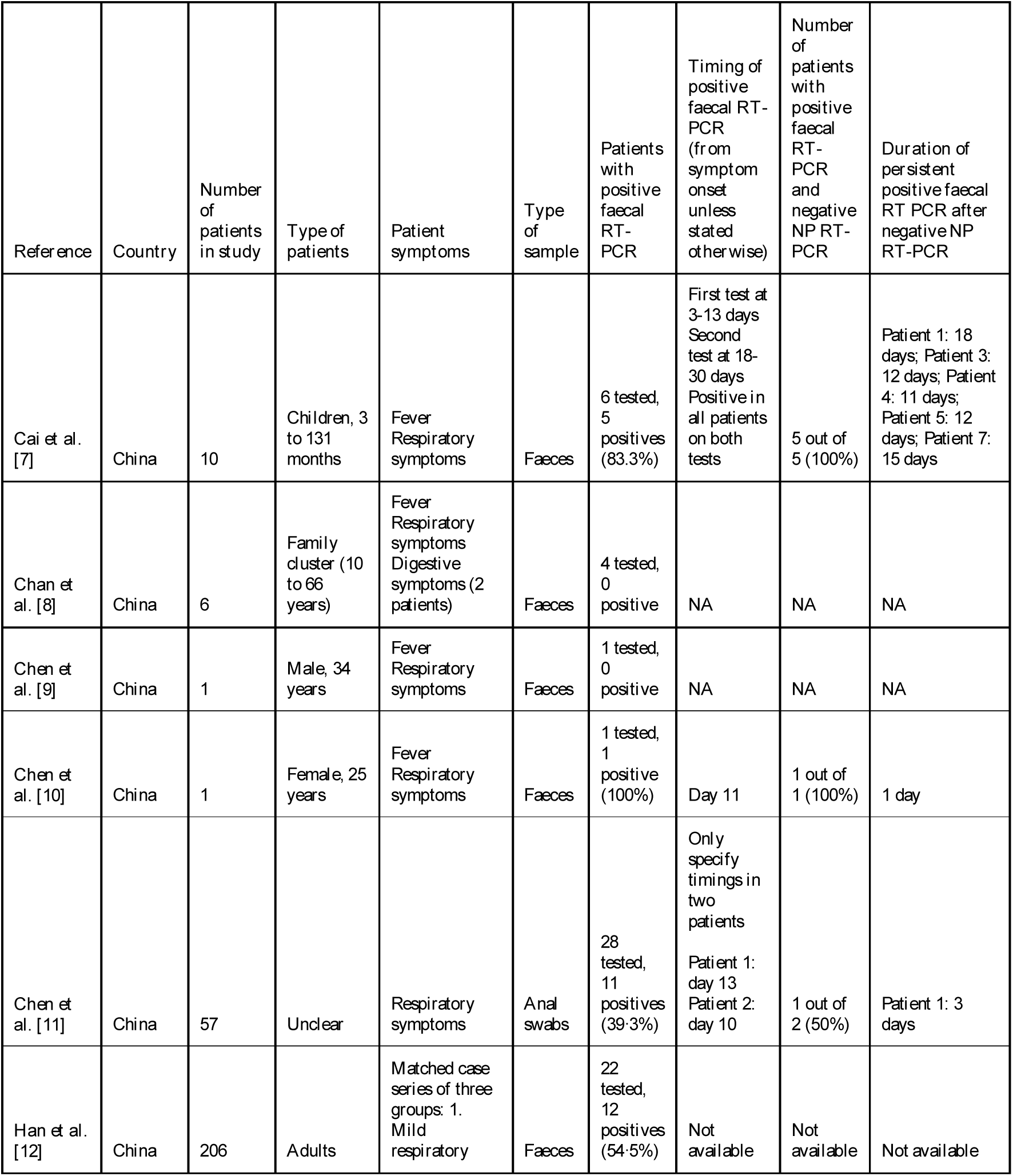
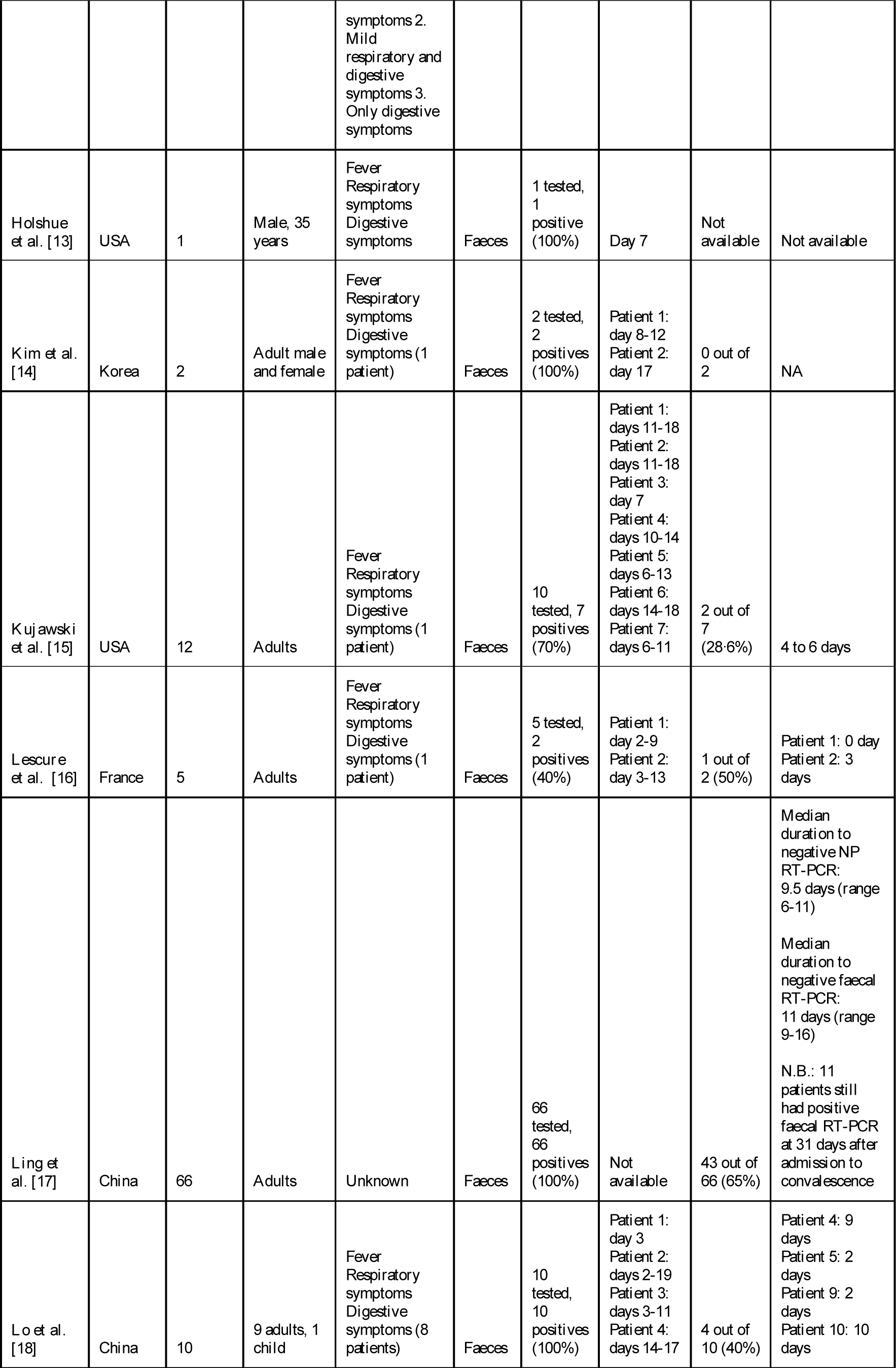
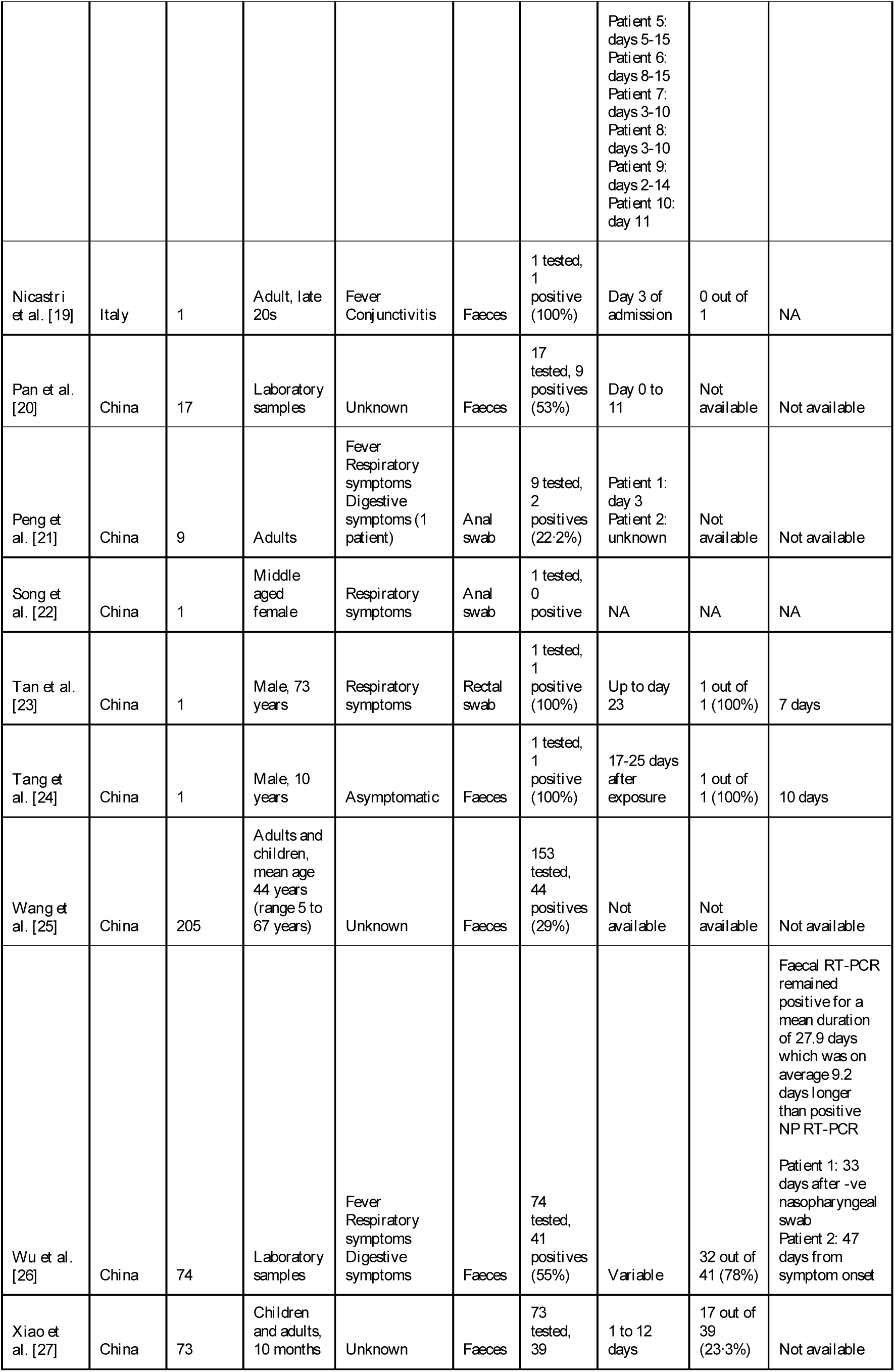
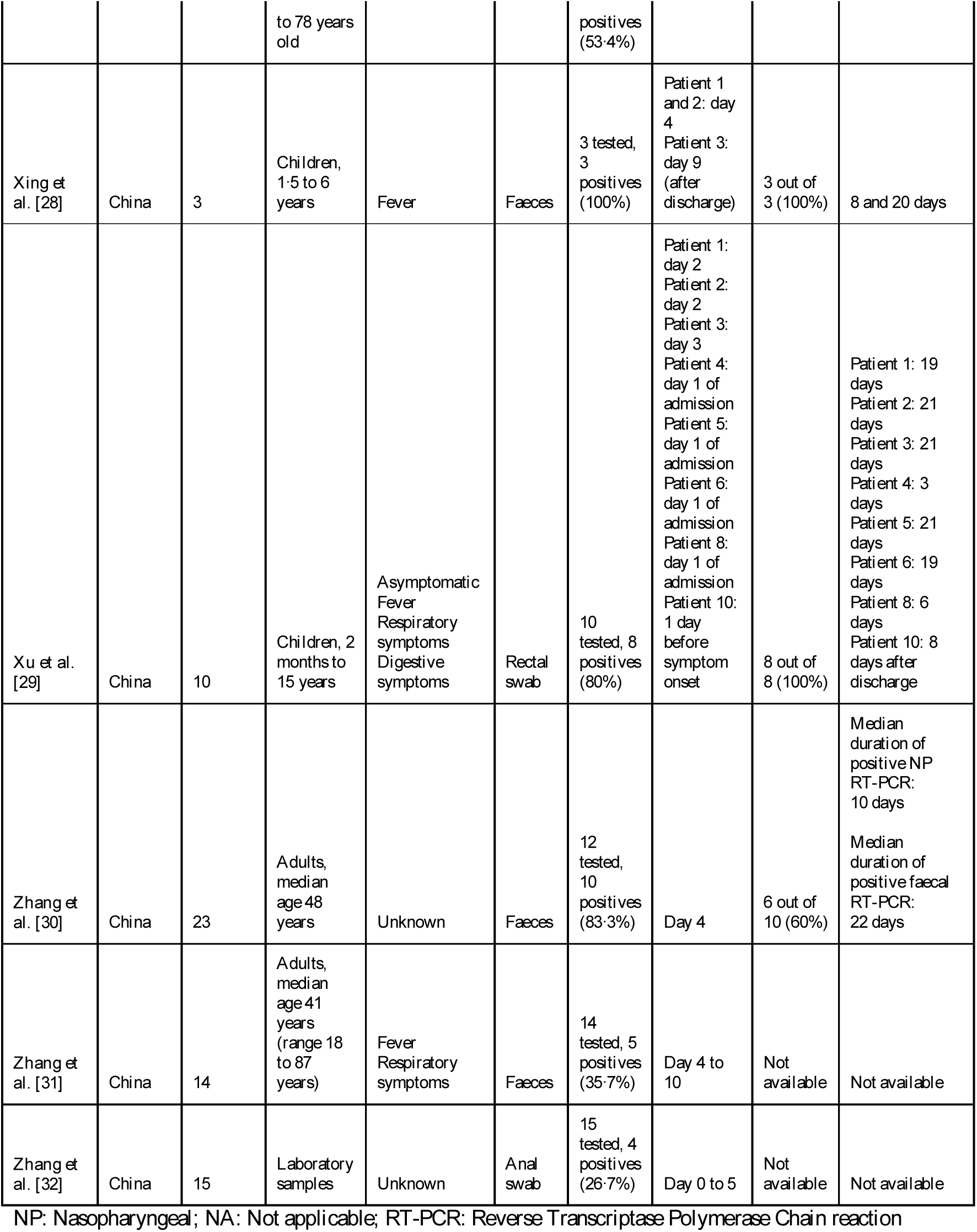

**Overview of data extracted from studies included in review [7–32]**

